# Genetic screening for *TLR7* variants in young and previously healthy men with severe COVID-19: a case series

**DOI:** 10.1101/2021.03.14.21252289

**Authors:** Xavier Solanich, Gardenia Vargas-Parra, Caspar I. van der Made, Annet Simons, Janneke Schuurs-Hoeijmakers, Arnau Antolí, Jesús del Valle, Gemma Rocamora-Blanch, Fernando Setién, Manel Esteller, Antoni Riera-Mestre, Joan Sabater-Riera, Gabriel Capellá, Frank L. van de Veerdonk, Ben van der Hoven, Xavier Corbella, Alexander Hoischen, Conxi Lázaro

## Abstract

Advanced age, male sex and chronic comorbidities are associated with severe COVID-19. However, these general risk factors cannot explain why critical illness occurs in young and apparently healthy individuals. In the past months, several publications have identified susceptibility loci and genes using comprehensive GWAS studies or genome, exome or candidate genes analysis. A recent study reported rare, loss-of-function *TLR7* variants in otherwise healthy young brother pairs from two families with severe COVID-19. We aimed to prospectively study the prevalence of rare X-chromosomal *TLR7* genetic variants in our cohort of young male patients with severe COVID-19. We recruited 13 patients ≤50 years who had no risk factors known to be associated with severe disease. We studied the entire *TLR7* coding region and identified two missense variants (p.Asn215Ser, c.644A>G and p.Trp933Arg, c.2797T>C) in two out of 13 cases (15.4%). These variants were not previously reported in population control databases (gnomAD) and were predicted to be damaging by all *in silico* predictors. The male index patients were between 25 and 30 years old and had no apparent comorbidities. The *TLR7* p.Asn215Ser co-segregated in 2 first-degree relatives severely affected by COVID-19, in a younger previously healthy the variant was found in hemizygous state, and in an older than 60 was in heterozygous state. No family members were available for testing the segregation of the p.Trp933Arg variant. These results further support that susceptibility to severe COVID-19 could be determined by inherited rare genetic variants in *TLR7*. Understanding the causes and mechanisms of life-threatening COVID-19 is crucial and could lead to novel preventive and therapeutic options. This study supports a rationale for the genetic screening for *TLR7* variants in young men with severe COVID-19 in the absence of other relevant risk factors. A diagnosis of TLR7 deficiency could not only inform on treatment options for the patient, but it also enables for pre-symptomatic testing of at-risk male relatives with the possibility of instituting early preventive and therapeutic interventions.

## INTRODUCTION

A proportion of patients with COVID-19 evolve to fatal lung injury and multi-organ failure due to systemic host-immune inflammatory processes triggered by the viral infection [1]. Advanced age, male sex and chronic disease such as diabetes and obesity are common in patients with more severe forms of COVID-19 [2-4]. However, these risk factors cannot explain why critical disease also occurs in younger (below 50 years of age) and apparently healthy individuals.

In the past months, several publications identified susceptibility loci and genes using comprehensive GWAS studies or genome, exome or candidate genes analysis. Hence, human genetic loci associated with a higher viral binding and entry comprised the *ABO* blood group locus [5-7], *ACE2* and *TMPRSS2* [8]. Genetic loci reported to predispose to higher severity include the 3p21.31 gene cluster [5], HLA-B*46:01 and HLA-B*15:03 [9], *APOE* [10], *IFITM3* [11], as well as several genes encoding for members of the type I/III interferon (IFN) pathway [12] including the Toll-Like Receptor 7 (*TLR7*) gene [13]. The Barcelona research group collaborated with a large genetic sequencing effort to define host risk factors to severe SARS-CoV-2 infection, analyzing exome or genome sequences from 659 patients with severe COVID-19 for rare pathogenic variants that could be associated with life-threatening disease [12]. This collaborative study was focused on the type I IFN pathway and analyzed 13 candidate genes (*TLR3, IRF7, IRF9, TICAM1/TRIF, UNC93B1, TRAF3, TBK1, IRF3, NEMO/IKBKG, IFNAR1, IFNAR2, STAT1* and *STAT2*) that have previously been linked with susceptibility to other viral infections. Loss of function variants were identified in 3.5% (23/659) of cases. In addition, a study of 156 Italian and Spanish <60 year-old patients with severe COVID-19, *TLR7* rare, but not entirely unique, missense variants were found in almost 4% and in none of the 122 oligo- or asymptomatic controls. Expression profiles of *TLR7* and type 1 IFN-related genes were studied in imiquimod-treated-PBMCs carrying 4 different variants (p.(Val219Ile), p.(Ser301Pro), p.(Ala1032Thr), p.(His630Tyr)) and the authors found a significant decrease of IRF7 and IFN-γ mRNA levels compared with healthy controls [Fallerini, preprint medRxiv 2020].

Here we describe two index patients with rare, putatively deleterious germline variants in the X-chromosomal *TLR7* gene. This finding reinforces the notion that TLR7 plays a critical role in the recognition of SARS-CoV-2 and the initiation of an early immune response to clear the virus and prevent the development of severe COVID-19. Our findings furthermore support the idea that, in some male patients, severe COVID-19 could be determined by rare *TLR7* variants and genetic screening may be appropriate in young, severely affected men without comorbidities predisposing to severe disease.

## METHODS AND RESULTS

This is a joint study performed at the Hospital Universitari de Bellvitge – IDIBELL, L’Hospitalet de Llobregat, Barcelona, Spain; and the Radboud University Medical Center, Nijmegen, The Netherlands and the Erasmus Medical Center, Rotterdam, The Netherlands.

### The Barcelona cases

From March to July 2020, researchers from Hospital Universitari de Bellvitge - IDIBELL prospectively collected biological samples from young patients without comorbidities related with severe COVID-19. Selection criteria were: 1) patients aged between 18 and 50 years old; 2) absence of known comorbidities associated with most severe forms of COVID-19; and 3) SARS-CoV-2 related lung injury requiring high flow oxygen devices or mechanical ventilation. Ten male patients fulfilled all selection criteria (Table 1). Eight patients (patients 1-8) were evaluated also by the COVID Human Genetic Effort where no pathogenic variants were detected in any of the 13 type I IFN pathway genes studied [12], none of those eight Barcelona patients carried a rare *TLR7* variant.

**Table 1.**
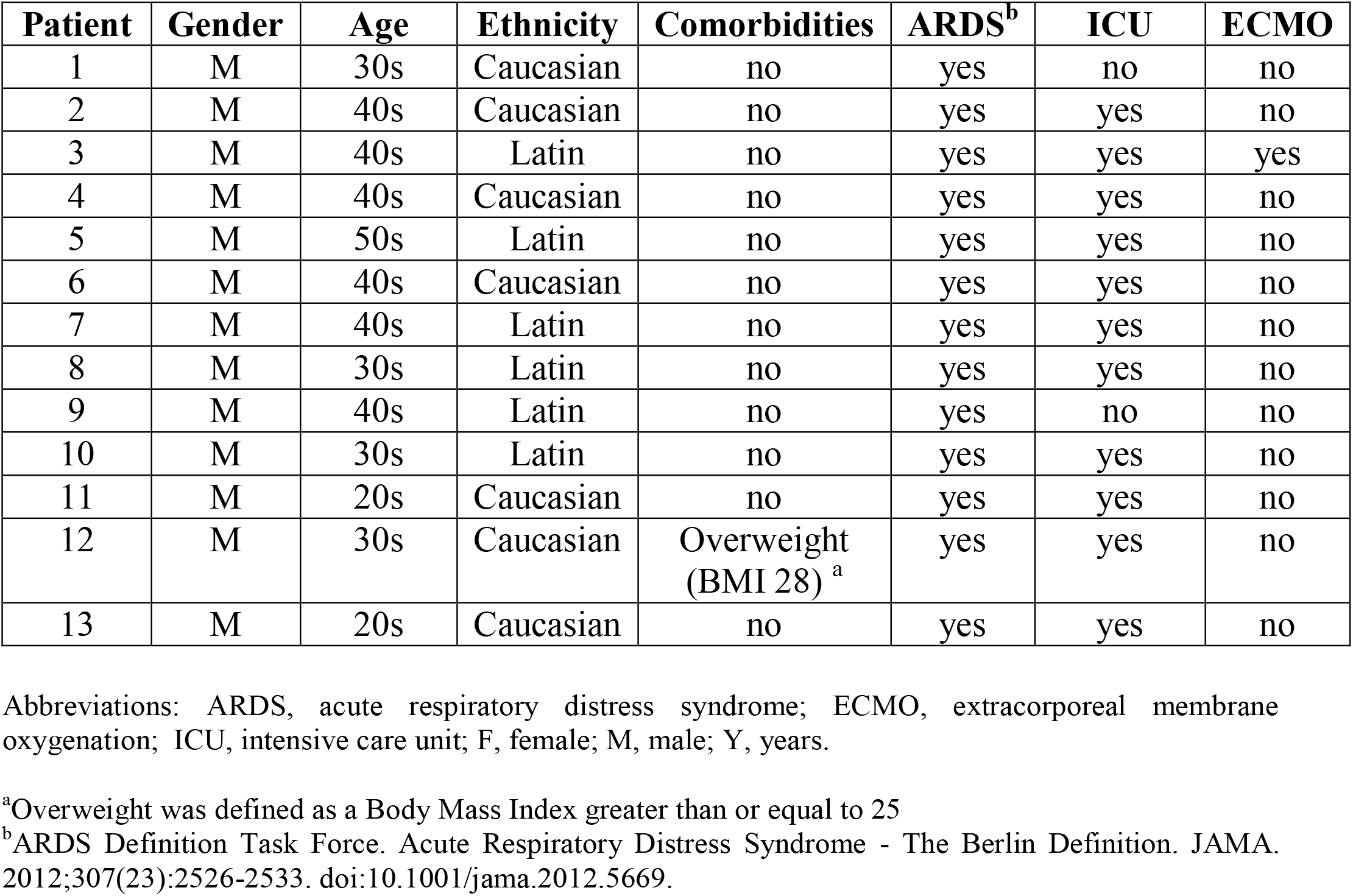
Demographic and Clinical Findings of Investigated Patients.

Informed consent was obtained from all patients and relatives, and the IDIBELL Research Ethics Committee approved this study (PR152/20). Demographic, epidemiological, laboratory and clinical data were collected. Treatments specifically used to treat COVID-19 at any time during admission were also documented.

DNA was isolated from total blood either using a Maxwell instrument RSC (Promega, Madison, WI, USA) or QIAGEN Flexigene DNA kit (Qiagen, Germany). Nine PCR primer pairs (Sigma-Aldrich, MO, USA) were designed to cover the whole coding region of *TLR7* gene. PCR was performed using DreamTaq MasterMix (ThermoFisher Scientific, Waltham, MA, USA), products were purified using EXO-SAP (New England Biolabs) and sequenced using the BigDye Terminator v.3.1 Sequencing Kit (Applied Biosystems, CA, USA) in an ABI Prism 3730 XL Genetic Analyzer (Applied Biosystems CA, USA). Primers and PCR conditions were available upon request. Mutation Surveyor software was used to detect variants and nomenclature was given according to HGVS guidelines. All variants identified were submitted to Alamut Software Suite v2.15.0 (Interactive Biosoftware) to retrieve population frequency and *in silico* prediction data.

### The Dutch cases

At the Radboud University Medical Center in Nijmegen and the Erasmus Medical Center in Rotterdam, the Netherlands, patients were screened prospectively in a clinical setting prospectively from December 2020 to February 2021 with the following criteria: 1) males aged below 40 years of age; 2) absence of comorbidities known to be associated with severe COVID-19 and 3) PCR-confirmed SARS-CoV-2 infection requiring high-flow oxygen therapy or ICU admission. A total of 3 patients (patients 11-13) fulfilled these inclusion criteria and underwent clinical Sanger sequencing (patient 11 and 12) or rapid whole-exome sequencing (patient 13) to specifically assess genetic variants in *TLR7*. Written informed consent was also obtained from patient 13 whose clinical data has been included in this study. Rapid whole-exome sequencing was performed similar to previous reports [13]. Sanger sequencing was done according to standard diagnostic procedures at the Department of Human Genetics, Radboud University Medical Center, protocols and primers sequences are available upon request.

### *TLR7* Sequencing Results

A total of 13 patients were included, with an average age of 37.85 (SD 9.026) years old. Putative deleterious *TLR7* variants were identified in two patients (patient 10 and 13). Both variants, which included a *TLR7* c.644A>G, p.(Asn215Ser) missense variant in patient 10 and a c.2797T>C p.(Trp933Arg) missense variant in patient 13, were not previously reported in our in-house database nor in the population database gnomAD [14]. Moreover, the Asn215Ser variant affects a highly conserved nucleotide and amino acid in the TLR7 leucine-rich region domain and it is predicted damaging or possibly damaging by *in silico* software. The Trp933Arg variant is also located at an evolutionarily highly conserved position within the TIR domain, important for downstream signaling via adapter proteins, and is considered deleterious by the *in silico* effect predictors. These variants and other previously reported variants in COVID-19 patients are shown schematically in Figure 1.

**Figure 1.**
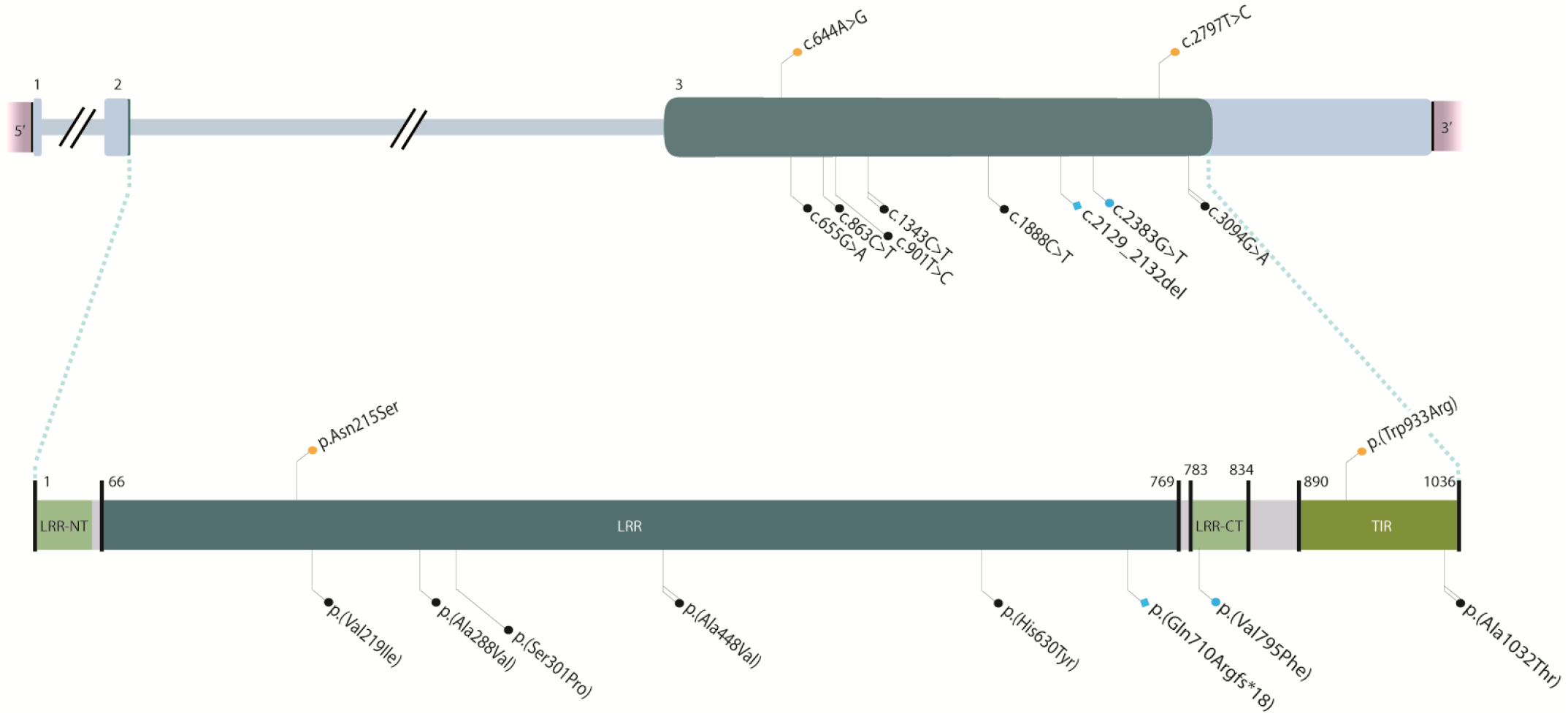
Schematic representation of *TLR7* variants reported to date in severely affected COVID-19 cases. Variants in cDNA (top) and protein (bottom). Color code: orange, variants found in the present series; blue, previously reported in van der Made, 2020; black, previously reported in Fallerini, 2020. Shape code: circle, missense variants; square, frameshift variants. Line code: single, reported in one case; double, reported in 2 cases.

### *TLR7* patients’ characteristics

Patient 10 was of Latin origin man in his 30s with no general risk factors predisposing to severe COVID-19. He developed pneumonia with bilateral consolidations on a computed tomography (CT) scan and fulfilled the criteria of acute respiratory distress syndrome (ARDS) secondary to PCR-proven COVID-19 (Table 2). The patient received antiviral treatment with remdesivir and immunosuppressive therapy with dexamethasone. Due to respiratory insufficiency, the patient was intubated and admitted at the ICU. The patient could be successfully extubated after 4 days of mechanical ventilation support and was discharged from ICU after 6 days. After the identification of the *TLR7* c.644A>G variant in the patient, segregation analysis confirmed segregation in both 2 first-degree relatives.. The younger relative (<30 years)was previously healthy without any comorbidity but, similarly to the index case, also contracted severe COVID-19, requiring mechanical ventilation and ICU admission at another hospital (Table 2). The brother pair of this family therefore represents the third pair of brothers with severe COVID-19, following the initial report [13]. Their older than 60 years relative suffered from obesity, dyslipidemia, type 2 diabetes and hypertension, and was also admitted at ICU due to critical respiratory failure caused by COVID-19. She was discharged from the ICU 16 days after admission. Main demographic, clinical, laboratory, and radiological findings of the three relatives are summarized in Table 2.

**Table 2.** Demographic, clinical, laboratory, and radiological findings of investigated patients.

|  | Patient 10 | Patient 10 –<br>First-degree<br>relative | Patient 10 –<br>First-degree<br>relative | Patient 13 | Reference<br>ranges |
| --- | --- | --- | --- | --- | --- |
| **Demographic characteristics** |  |  |  |  |  |
| Age, decade | 30s | 20s | 60s | 20s |  |
| Sex | Male | Male | Female | Male |  |
| Medical history | None | None | Obesity,<br>dyslipidemia,<br>hypertension,<br>type 2 diabetes, | Vasovagal<br>syncope |  |
| **Clinical characteristics at presentation** |  |  |  |  |  |
| Time from symptom onset to hospitalization, d | 7 | 6 | 12 | 2 |  |
| Symptoms at disease onset | Dyspnea, cough,<br>fever, myalgia | Dyspnea, cough,<br>fever, headache | Dyspnea, cough,<br>fever, myalgia | Dyspnea,<br>cough, fever,<br>respiratory<br>arrest |  |
| Imaging features (CT scan) | Bilateral<br>pulmonary<br>consolidations | Bilateral<br>pulmonary<br>consolidations | Bilateral<br>pulmonary<br>consolidations | Multiple<br>ground glass<br>opacities and<br>consolidations<br>in all lung<br>segments |  |
| **ICU admission** |  |  |  |  |  |
| Time from symptom onset to ICU admission, d | 8 | 7 | 12 | 7 |  |
| Medical reason for ICU admission | Respiratory<br>insufficiency | Respiratory<br>insufficiency | Respiratory<br>insufficiency | Respiratory<br>failure,<br>respiratory<br>arrest,<br>resuscitation<br>at home. |  |
| Disease severity status on admission, SOFA<br>score<sup>a</sup> | 3 | 3 | 4 | 6 |  |
| **Laboratory findings at ICU admission** |  |  |  |  |  |
| **Chemistry** |  |  |  |  |  |
| Alanine aminotransferase, U/L | 135 | 14 | 23.5 | 41 | <40 |
| Albumin, g/L | 37 | 31 | 28.1 | 23 | 35 - 52 |
| Alkaline phosphatase, U/L | 131 | 66 | 84.0 | 222 | ≤ 129 |
| Aspartate aminotransferase, U/L | 92 | 22 | 73.8 | 37 | ≤ 39 |
| Cardiac troponin, high sensitivity, ng/L | NA | 7 | NA | NA | ≤ 13 |
| Creatine kinase, U/L | 51 | 35 | 180 | NA | ≤ 189 |
| Creatinine, μmol/L | 60 | 57 | 57.1 | 84 | 44-97 |
| eGFR, mL/min/1.73 m<sup>2</sup> | >90 | >90 | >90 | >90 | >90 |
| γ-Glutamyltransferase, U/L | 243 | 27 | 34.8 | 263 | ≤ 70 |
| Lactate dehydrogenase, U/L | 381 | 432 | 1088.4 | 201 | <250 |
| **Blood count** |  |  |  |  |  |
| Hemoglobin, g/L | 112 | 114 | 110 | 121 | 130 - 165 |
| Lymphocyte count, ×10<sup>9</sup>/L | 1.8 | 1.47 | 2.19 | 1.64 | 1.3-3.4 |
| White blood cell count, ×10<sup>9</sup>/L | 18 | 10.7 | 14.74 | 8.4 | 3.9-9.5 |
| Platelet count, ×10<sup>9</sup>/L | 385 | 408 | 416 | 369 | 149 - 303 |
| **Coagulation** |  |  |  |  |  |
| Activated partial thromboplastin time ratio | 0.95 | 0.98 | 1.00 | 37 | 0.8-1.2 |
| D-dimer, ng/mL | <250 | 463 | 2400 | 3660 | <250 |
| Fibrinogen, g/L | > 7 | > 7 | 5.5 | 4.2 | 2.76-4.71 |
| Prothrombin time ratio | 1.38 | 1.32 | 1.10 | NA | 0.8-1.2 |
| **Inflammatory markers** |  |  |  |  |  |
| C-reactive protein, mg/L | 346.6 | 267 | 203.98 | 196 | <3 |
| Ferritin, µg/L | 1957.6 | 920 | 384.9 | 845 | 30 - 400 |
| Procalcitonin, µg/L | 0.28 | NA | 0.1 | 4.31 | <0.5 |
| IL-6, ng/L | 48.4 | 1.5 | 24 | NA | ≤ 6.9 |
| Secondary complications | None reported | catheter-related bloodstream infection | None reported | Small ventral pneumothorax at admission after resuscitation at home. Bilateral subsegmental pulmonary embolisms. |  |
| Duration of viral shedding after COVID-19 onset (positive SARS-CoV-2 PCR), d | Positive at admission, no follow-up measurement | Positive at admission, no follow-up measurement | Positive at admission, no follow-up measurement | Positive before admission, PCR negative at day 29 |  |
| Duration of ventilatory support, d | 4 | 10 | 7 | 24 days (ongoing) |  |
| Duration of ICU stay, d | 6 | 12 | 16 | 24 days (ongoing) |  |
| **Follow-up** |  |  |  |  |  |
| Time from ICU discharge to hospital discharge, d | 3 | 11 | 5 | NA |  |
| Complications during follow-up period | None reported | None reported | None reported | NA |  |
| Treatments | R, D | H, L/R, MP, I, T | R, D, T | D |  |

Abbreviations: COVID-19, coronavirus disease 2019; CT, computed tomography; ICU, intensive care unit; eGFR, estimated glomerular filtration rate; NA, not assessed; PCR, polymerase chain reaction; SARS-CoV-2; severe acute respiratory syndrome coronavirus 2; SOFA, Sequential Organ Failure Assessment. The treatments were administered to the 3 patients as follows: D, Dexamethasone 6 mg / day intravenously for 10 days; R, Remdesivir 200mg orally the first day and 100mg every day the next 4 days; T, Tocilizumab 600 mg single dose intravenously; L/R, Lopinavir 400mg/Ritonavir 100mg orally every 12 hours for 3d; H, Hydroxychloroquine orally 400mg every 12 hours the first day and 200mg every 12 hours the next 10 days; I, Interferon β1b 0.25mg every 48 hours subcutaneously for 3 days.
<sup>a</sup> The SOFA score is calculated using 6 systems: respiratory, coagulation, hepatic, cardiovascular, central nervous, and kidney. Scores range from 0 for normal function to 4 for most abnormal and are summed for a final range of 0 to 24. An initial score of 2 to 3 is associated with 6 % mortality; an initial score of 4 to 5 is associated with 20 % mortality.

Patient 13 was a Caucasian male in his 20s without previous medical history or comorbidities. The patient complained of progressive dyspnea and shortness of breath and due to rapid clinical deterioration and respiratory insufficiency, he was intubated by the medical emergency team at his home and subsequently hospitalized in ICU at a peripheral hospital. A CT-scan showed multiple diffuse ground-glass opacities and consolidations in all lung segments, meeting the criteria for ARDS. Treatment consisted of mechanical ventilation with prone positioning, intravenous dexamethasone, and the antibiotics ceftriaxone and ciprofloxacin. However, the patient’s condition further deteriorated and he was referred to the Erasmus Medical Center for possible ECMO treatment. However, with continuing prone positioning he gradually improved before ECMO was required. A repeated CT scan also showed subsegmental pulmonary embolisms for which intravenous heparin was started. In the following weeks, the patient gradually recovered and to date remains in the ICU after a total of 26 days. He has started physical therapy and is weaning of the ventilator.

The patients’ whole family contracted COVID-19 at the time the patient developed symptoms, including his brother, who had only minor symptoms. Variant segregation analysis is still pending.

## DISCUSSION

In July 2020, rare, deleterious, germline variants in the X-chromosomal *TLR7* gene were reported in young and, otherwise, healthy males with severe COVID-19. In these two brother pairs, rapid whole-exome sequencing identified both a maternally inherited 4-nucleotide deletion (c.2129_2132del; p.(Gln710Argfs*18)) and a missense variant (c.2383G>T; p.(Val795Phe)). Both variants were associated with impaired type I and II IFN responses, showing its importance in the COVID-19 pathogenesis [13]. Since *TLR7* was not one of the genes initially evaluated by the COVID Human Genetic Effort, our Barcelona team decided to perform a *TLR7* Sanger sequencing analysis in their case series consisting of 10 healthy young males with severe COVID-19. We identified one patient with a new missense variant (c.644A>G; p.(Asn215Ser)) labelled as damaging by *in silico* predictors. In addition, three Dutch cases were added to this series who underwent genetic screening in a clinical setting, leading to the identification of another novel, unique missense variant (c.2797T>C; p.(Trp933Arg)) located in the highly conserved TIR domain. While we cannot unequivocally prove the pathogenicity of the variants identified, the odds of identifying two private missense variants in just 13 cases is very unlikely a chance finding, considering the extremely rare prevalence of *TLR7* variants in gnomAD.

The finding of these two variants in a total of 13 screened patients (15.4%) led us to consider that *TLR7* variants could be a relatively common cause predisposing to severe COVID-19 in this subset of young, male patients with absence of general risk factors for severe disease. However, it should be noted that complete *TLR7* deficiency is estimated to be extremely rare. Endosomal TLRs (TLR3, TLR7, TLR8, and TLR9) have evolved under strong purifying selection. This selective regime ensures the conservation of particularly important proteins and indicates that these receptors play an essential, non-redundant biological role in host survival [15-16]. Therefore, rare *TLR7* non-synonymous variants are unlikely to be an explanation for severe COVID-19 in the general population. It is intriguing to speculate that some, very rare variants compromising essential functional domains in TLR7 lead to a complete deficiency [13], while other, relatively less rare variants may lead only to a partial TLR7 deficiency, and therefore may impact a larger group of individuals, but exert a lower relative risk to develop severe COVID-19. It is therefore interesting, that these latter variants have been also be identified in individuals of elevated age and also compromise TLR7 function [Fallerini, preprint MedRxiv 2020].

Common low effect size genetic variants in *TLR7* have been proposed as a possible explanation of the male sex bias in COVID-19 severity because of its localization on the X chromosome and well-established function in innate immunity [17]. Interestingly, circulating 25-hydroxy vitamin D (25OHD) levels decline with age [18] and may be accompanied by a lower expression and defective function of TLR7 [19]. Accordingly, TLR7 function could be modified by genetic but also epidemiological factors and certain comorbidities. So it is possible, but only speculative, that the addition of several factors that modify TLR7 function may be a common cause of progression to the most severe stages of COVID-19 in male. The same speculative hypothesis could be applicable to other genes involved in immune response regulation after SARS-CoV-2 infection [4,20].

Specific genetic variants that confer a higher risk to biological agents such as HIV, malaria and tuberculosis have been identified through NGS and genome wide association studies (GWAS) [21]. These host DNA variants now serve as biomarkers that can be used for early diagnosis and prophylaxis, and allow the identification of possible molecular targets for treatment. This also applies to susceptibility to SARS-CoV-2 that could be determined by genes related to viral binding and entry, as well as genes related with immune response to the SARS-CoV-2 [4]. The small cohort study presented here, with an unexpectedly high yield (2/13) encourages that a screen for *TLR7* rare variants in severely affected men may be useful. While also elderly individuals may carry rare *TLR7* variants those individuals may be more difficult to identify; we therefore suggest the following preliminary screening criteria: young men (<50 year of age); previously healthy; suffering from severe COVID-19 in addition affected young brother pairs – as well as pedigrees suggestive for X-linked segregation - should be further prioritized.

Unfortunately, there is still a paucity of therapies that has been proven effective and safe in the treatment of COVID-19, other than supportive care and dexamethasone [22]. Moreover, even less is known about how patients with pathogenic variants in type I IFN pathway genes should be treated. Studying the pathways critically involved in the pathogenesis of severe COVID-19 could lead to novel preventative and therapeutic options to treat such patients and also their at-risk male relatives. In this respect, there would be a strong argument to offer hemizygous *TLR7* deficient males that have not had COVID-19 direct access to vaccination as effective preventative measure, similar to other patients with primary immunodeficiencies.

In summary, host genetics should be evaluated in young and apparently healthy individuals with life-threatening COVID-19. We describe two new germline putative deleterious variants in the X chromosomal *TLR7* gene. To know host DNA variants related with SARS-CoV-2 susceptibility may help physicians to identify and treat those patients at higher risk to develop severe COVID-19. In this way, their relatives at risk could be offered options for pre-symptomatic tests in order to early establish preventive measures. Further studies are needed to determine the pathogenicity of the variants reported here, as well as the prevalence of pathogenic variants in *TLR7* in larger cohorts of young, healthy male patients severely affected by SARS-CoV-2. Finally, the contribution of genetic variation in *TLR7* in the population of older healthy male patients should also be assessed.

## Data Availability

The raw data supporting the conclusions of this article will be made available by the authors, without undue reservation.

## Ethics statement

Ethical approval for the study was obtained from the Hospital Universitari de Bellvitge – IDIBELL (L’Hospitalet de Llobregat, Barcelona, Spain) Research Ethics Committee (approval number PR152/20). The patients/participants provided their written informed consent to participate in this study. Patients from the Netherlands gave consent for diagnostic testing of *TLR7* and/or whole exome sequencing, patient 13 gave written consent for publication of his clinical data.

## Author contributions

XS, GVP and CIvdM contributed equally to this work. XS, AH and CL devised the study. GC, CIvdM, ARM, FS, ME and XC provided input on the study design. AA, BvdH, JSH, FvdV and GRB assisted in patient management. GVP, AS and JdV designed and performed the sequence analysis. XS, GVP, CIvdM, AH and CL had full access to all data and take responsibility for the integrity and the accuracy of the data. XS, GVP, CvDM, AH and CL drafted the manuscript. All authors revised and approved the final manuscript.

## Acknowledgments

We sincerely thank the patients and their families for their participation. With the support of the Departament de Salut de la Generalitat de Catalunya. We thank CERCA Programme / Generalitat de Catalunya for institutional support. We also thank the Radboud Genomics Technology Center for their technical support.

